# Plasma phosphorylated tau 181 predicts amyloid status and conversion to dementia stage dependent on renal function

**DOI:** 10.1101/2022.09.13.22279818

**Authors:** Sylvain Lehmann, Susanna Schraen-Maschke, Jean-Sébastien Vidal, Constance Delaby, Frédéric Blanc, Claire Paquet, Bernadette Allinquant, Stéphanie Bombois, Audrey Gabelle, Olivier Hanon, the BALTAZAR study group.

## Abstract

**Objectives:** Plasma P-tau181 is an increasingly established diagnostic marker for Alzheimer’s disease (AD). Further validation in prospective cohorts is still needed, as well as the study of confounding factors that could influence its blood level.

**Methods:** This study is ancillary to the prospective multicenter BALTAZAR cohort that enrolled participants with mild cognitive impairment (MCI) who were examined for conversion to dementia for up to three years. Plasma Ptau-181 was measured using the ultrasensitive Quanterix HD-X assay.

**Results:** Among 476 MCI participants, 67% were Aβ+ at baseline and 30% developed dementia. Plasma P-tau181 was higher in the Aβ+ population (3.9 [SD 1.4] vs. 2.6 [SD 1.4] pg/mL) and in MCI that converted to dementia (3.8 [SD 1.5] vs. 2.9 [SD 1.4] pg/mL). The addition of plasma P-tau181 to a logistic regression model combining age, sex, APOEε4 status and mini mental state examination (MMSE) improved predictive performance (AUC 0.691 to 0.744 for conversion and 0.786 to 0.849 for Aβ+). The Kaplan–Meier curve of conversion to dementia, according to the tertiles of plasma P-tau181, revealed a significant predictive value (Log rank P<.0001) with a hazard ratio of 3.8 [95%CI=2.5-5.8]. In addition, patients with plasma P-Tau(181) ≤2.32 pg/mL had a conversion rate of less than 20% over a 3-year period. Using a linear regression approach, chronic kidney disease (CKD), creatinine and glomerular filtration rate (eGFR) were independently associated with plasma P-tau181 concentrations.

**Conclusions:** Plasma P-tau181 effectively detects Aβ+ status and conversion to dementia, confirming the value of this blood biomarker for the management of AD. However, renal function significantly modifies its levels and may thus induce diagnostic errors if not taken into account.

**What is already known on this topic:** The clinical use of plasma phosphorylated tau 181 (P-tau181) for Alzheimer’s disease is being considered but further validation and study of confounding factors are still needed.

**What this study adds:** In our large prospective cohort, P-tau181 predicts brain amyloidopathy and conversion to dementia in patients with mild cognitive impairment, but renal function significantly alters plasma levels and thus may induce diagnostic errors if not taken into account.

**How this study might affect research, practice or policy:** Our study suggests that measurement of creatinine or estimation of glomerular filtration rate, which are easy and standardizable ways to provide information on renal function, will contribute to optimal interpretation of plasma P-tau181 results in routine clinical practice.

## Introduction

Alzheimer’s disease (AD) accounts for 60% to 70% of dementia and is thus a major public health problem and socio-economic burden that is only increasing with the ageing of the population. For a long time, diagnostic efforts have been minimal, due to the lack of preventive measures or curative treatment. However, in recent years, it has been demonstrated that modifiable risk factors account for 40% of AD ^1^. Furthermore, many potential treatments are in the final stages of study^2^. Early diagnosis will be the key to further understanding of AD and successfully treating it.

The development of biological biomarkers, linked to amyloid and tau pathology, has greatly contributed to the understanding of the pre- and post-symptomatic AD “continuum”, in which, thanks to genetic forms, it has been shown that the disease was present from a biological point of view decades before its clinical appearance ^3^. This opens up therapeutic perspectives and allows us to use these biomarkers in early diagnosis and even in risk assessment. Their relevance has already been proven in cerebrospinal fluid (CSF) justifying their implementation in clinical routine^4 5^. Furthermore, the blood is now amenable to diagnostic assays, thanks to ultrasensitive techniques, including mass spectrometry.

The first analytes used in blood were amyloid peptides. Their levels in serum can detect amyloid positive (Aβ+) patients, as defined by amyloid PET or CSF analysis, and predict evolution of patients within the AD continuum, including conversion of patients, with mild cognitive impairment (MCI), to dementia ^6^. While the detection of total tau in blood was not so discriminating ^7^, detection of its phosphorylated forms at position threonine 181 was a real breakthrough ^8^. P-tau181 concentration is predictive of amyloid or tau PET findings and is significantly higher in AD and MCI compared to subjects without cognitive impairment or compared to other causes of dementia ^8-10^. Its prognostic value has also been established in some longitudinal studies ^11-13^ and its overall performance allow to consider a clinical application. To reach this milestone, we will need additional prospective data, *in vitro* diagnostic (IVD) certified kits and sufficient information on preanalytical stability ^14 15^. It will equally be paramount to identify any confounding factors that may alter clinical performance in routine use^14^.

In this work, we used the prospective multicenter BALTAZAR cohort ^16^ to confirm the ability of plasma P-tau181 to detect brain amyloidopathy and also to predict the conversion of MCI patients to dementia stage. As plasma biomarkers are soon to be implemented in routine practice, we also investigated potential confounding factors that must be considered to interpret the results adequately.

## Materials and Methods

### Study population

The BALTAZAR (Biomarker of AmyLoid pepTide and AlZheimer’s diseAse Risk) study is a multicenter prospective cohort study (ClinicalTrials.gov Identifier #NCT01315639) that enrolled patients with MCI or AD, according to a previously described protocol ^16^. All participants had clinical, neuropsychological, brain MRI and biological assessments (see below). Right and left hippocampal volume was obtain for each participant using automatic segmentation of the hippocampus. The hippocampal volume was normalized using the following calculation: hippocampal volume/total brain volume × mean total brain volume. CSF samples were collected only in accepting participants. APOE was genotyped in a single centralized laboratory. MCI subjects were selected according to the Petersen’ criteria ^17^ and then they were dichotomized into amnestic (aMCI) and non-amnestic (naMCI) phenotypes, based on the presence of memory impairment on the free and cued selective reminding test related to age, sex and educational level. The characteristic of the aMCI/naMCI population is fully described elsewhere ^16^. Patients had visits every six months for three years. MCI participants were reassessed for conversion to dementia at each visit by the clinician ^6^. The progression from MCI to dementia was defined by evaluating the following parameters: (i) decline in cognitive function (measured by changes from the baseline in scores of the mini mental state examination [MMSE]), (ii) disability in activities of daily living (instrumental activities of daily living iADL>1) and (iii) clinical dementia rating sum of boxes (> 1). The conversions were reviewed by an adjudication committee.

In this study, we analyzed 476 available baseline plasma samples from patients with MCI diagnosis (365 aMCI and 111 naMCI).

### Biological biomarker measurements

To minimize pre-analytical and analytical problems, identical collection tubes were used across centers to collect plasma (EDTA BD Vacutainer K2E, ref 367 525, Becton Dickinson, USA) and for CSF (10mL polypropylene tube, ref 62.610.201, Sarstedt, Germany). Blood and CSF samples were collected on the same day. All aliquots were stored in the same low-binding Eppendorf® LoBind microtubes (Eppendorf, ref 022431064, Hamburg, Germany). Baseline blood samples were used to measure fasting glycemia, cholesterol (total, HDL, LDL), prealbumin, albumin, creatinine ^16^. Estimated glomerular filtration rate (eGFR), based on creatinine, age and sex, was computed using the CKD-Epidemiology Collaboration (CKD-EPI) equation ^18^. CSF biomarkers were measured in a single centralized laboratory using commercially available Innotest® assays for tau and phosphorylated tau at position T181 (P-tau181) or Euroimmun® for amyloid peptides Aβ1-42 and Aβ1-40. Positive amyloid status (Aβ+) was defined, as previously, when the CSF Aβ1-42/Aβ1-40 ratio was below 0.1 ^19^.

Plasma P-tau181 was determined using a commercial P-tau181 assay kit (Quanterix, USA) based on ultrasensitive Simoa technology ^20^ on an HD-X analytical platform. All samples were 4-fold diluted with the provided dilution buffer to minimize matrix effects. After dilution, the lowest limit of detection was 0.019 pg/mL and the limit of quantification was of 0.085 pg/mL. Quality controls with low (QC 1 with mean concentration of 3.82 pg/mL) or high (QC 2 - 52.4 pg/mL) P-tau181 known concentration were provided in the kits. Inter-assay variation for QC 1 and QC 2 was low, with coefficient of variation (CV) of 7% and 5%, respectively. We also used two serum pools (average P-tau181 of 4.47 pg/mL and 2.81 pg/mL) as internal QCs run at the beginning and end of each sample plate. These had low inter-assay CV of 3% and 6%, respectively.

### Statistical analyses

General characteristics were analyzed in the whole MCI sample, according to MCI subtype (aMCI and naMCI), conversion to dementia, and to plasma P-tau181 tertile. Categorical variables are presented as percentages and counts (% (N)); continuous variables, as mean and standard deviation (M (SD)), or median [25-75 percentile], and comparisons were assessed by χ^2^ tests, T-tests, Mann-Whitney and analysis of variance (ANOVA, Kruskal-Wallis test). The relationship between conversion and plasma P-tau181 was assessed using regression models with age, sex, and baseline presence of APOE ε4 allele as covariables.

Kaplan Meier curves were drawn for conversion according to plasma P-tau181tertile and overall differences between tertiles was calculated by Log rank test. We also examined how plasma P-tau181 improved dementia risk prediction using logistic regression with age, sex, APOE ε4 and MMSE score at baseline and by calculating continuous net reclassification improvement (NRI) ^21^. Receiving Operator Characteristic (ROC) curves, using conversion as a dependent variable, were also used to compute for different factors. The corresponding areas under the curve (AUCs) were compared using the Delong method ^22^. Logistic regression model (enter model), Kaplan Meier and ROC curves were generated with MedCalc (20.111) software. In all analyses, the 2-sided α-level of 0.05 was used for significance testing.

## Results

### Characteristics of the MCI participants at baseline

Of the 539 MCI participants enrolled in the BALTAZAR study, 63 were excluded due to missing data or absence of plasma P-tau181 biomarkers. In this study, we analyzed, 476 MCI participants (mean age 77.7 [SD 5.5] years, 61.4% women) with 365 aMCI (77%) and 111 naMCI (23%) at baseline (Table 1, Suppl Table 1). Average MMSE score was 26.4 [SD 2.5] and 39.8% (n=185) were APOE ε4 carriers. During the clinical follow-up period of 6 to 36 months, 30% (n=144) of the MCI participants developed dementia, on average 14.6 [SD 8.2] months after the baseline visit and in 95% of the cases, they converted to clinically probable AD ^6^.

**Table 1:** Characteristics in the whole MCI population and between MCI participants who converted, or not, to dementia within 3 years.

| Patient characteristics | All MCI<br>N=476 | MCI Non-converters<br>N= 332 | MCI converters<br>N=144 | P | P\$ |
| --- | --- | --- | --- | --- | --- |
| Age (years) | 77.7 (5.5) | 77.3 (5.4) | 78.5 (5.7) | **.048** | **.0033** |
| Women (%) | 292 (61.4) | 205 (61.7) | 60.4 | .78 | .67 |
| BMI (kg/m<sup>2</sup>) | 25 (3.8) | 25.1 (3.8) | 24.8 (3.8) | .39 | .95 |
| MMSE (/30) | 26.4 (2.5) | 26.7 (2.5) | 25.6 (2.5) | **<.0001** | **.0002** |
| 1 or 2 APOE4 alleles | 185 (39.8) | 105 (31.6) | 80 (55.5) | **<.0001** | **<.0001** |
| Hippocampal volume (R+L) (cm<sup>3</sup>) | 4.55 (1.12) | 4.79 (1.05) | 4.01 (1.09) | **<.0001** | **<.0001** |
| CSF biomarkers* | All MCI | MCI Non-converters | MCI converters | P | P\$ |
| Aβ1-40 (pg/mL) | 7434 (2241) | 7458 (2298) | 7389 (2143) | .83 | .64 |
| Aβ1-42 (pg/mL) | 766 (384) | 857 (398) | 593 (288) | **<.0001** | **<.0001** |
| Aβ1-42/Aβ1-40 | 0.104 (0.045) | 0.116 (0.045) | 0.082 (0.036) | **<.0001** | **<.0001** |
| Tau (pg/mL) | 578 (254) | 376.1 (185.2) | 547 (223.4) | **<.0001** | **<.0001** |
| p-Tau181 (pg/mL) | 76.7 (30.2) | 58.58 (24.47) | 80.3 (33.58) | **<.0001** | **<.0001** |
| Blood biomarkers | All MCI | MCI Non-converters | MCI converters | P | P\$ |
| Fasting glycemia (mmol/L) | 5.37 (1.19) | 5.34 (1.21) | 5.45 (1.14) | .34 | .20 |
| Triglycerides (mmol/L) | 1.21 (0.59) | 1.2 (0.58) | 1.24 (0.6) | 0.54 | .82 |
| Cholesterol total (mmol/L) | 5.5 (1.16) | 5.48 (1.2) | 5.53 (1.07) | .67 | .91 |
| Cholesterol HDL (mmol/L) | 1.74 (0.52) | 1.75 (0.53) | 1.72 (0.48) | .66 | .98 |
| Cholesterol LDL (mmol/L) | 3.2 (1) | 3.19 (1.01) | 3.24 (0.98) | .65 | .90 |
| Prealbumin (mg/dl) | 27.6 (5.4) | 28.1 (6.2) | 27.7 (5.8) | .53 | .52 |
| Albumin (g/L) | 40.3 (3.9) | 40.5 (3.5) | 39.8 (4.5) | .088 | .11 |
| Creatinine (μmol/L) | 78.2 (21.3) | 79.4 (22.2) | 73.7 (13.6) | .10 | .82 |
| eGFR (mL/min/1.73m<sup>2</sup>) | 76.9 (14.7) | 77.0 (14.8) | 76.7 (14.7) | .85 | .80 |
| Plasma P-tau181(pg/mL) | 3.19 (1.49) | 2.9 (1.4) | 3.8 (1.5) | **<.0001** | **<.0001** |

Abbreviations: MCI: mild cognitive impairment; APOE: apolipoprotein E; BMI, body mass index; eGFR, estimated glomerular filtration rate; MMSE, Mini-Mental State Examination; R+L, right+left.
P: Comparison between the three groups, by ANOVA or $\chi^2$ ; P\$: comparison between the three groups by linear regression adjusted for age, sex, and the presence of the APOE ε4 allele; % (number) were used to describe categorical variable, mean ± standard deviation for continuous variables.
\* CSF biomarkers were available in 140 and 74 MCI non-converters and converters, respectively.

**Table 2:** Characteristics in the whole population and between Aβ- and + patients.

| Patient characteristics | All<br>N=214 | Aβ-<br>N= 97 | Aβ+<br>N=117 | P | P\$ |
| --- | --- | --- | --- | --- | --- |
| Age (years) | 77.4 (5.6) | 76.6 (5.1) | 78 (5.9) | .073 | **.0122** |
| Women (%) | 127 (59.3) | 53 (54.6) | 74(64.9) | .20 | .36 |
| BMI (kg/m<sup>2</sup>) | 24.7 (3.7) | 25.4 (3.7) | 24.2 (3.6) | **.024** | .26 |
| MMSE (/30) | 26.4 (2.4) | 27.1 (2) | 25.8 (2.5) | **<.0001** | **0.0007** |
| 1 or 2 APOE4 alleles | 78 (36.4) | 15 (15.5) | 63 (57.3) | **<.0001** | **<.0001** |
| Hippocampal volume (R+L) (cm<sup>3</sup>) | 4.56 (1.09) | 4.64 (1.23) | 4.5 (0.96) | .41 | .91 |
| CSF biomarkers* | All | Aβ- | Aβ+ | P | P\$ |
| Aβ1-40 (pg/mL) | 7434 (2241) | 7421 (1896) | 7446 (2499) | .93 | .62 |
| Aβ1-42 (pg/mL) | 766 (385) | 1095 (287) | 494 (196) | **<.0001** | **<.0001** |
| Aβ1-42/Aβ1-40 | 0.104 (0.045) | 0.149 (0.022) | 0.068 (0.018) | **<.0001** | **<.0001** |
| Tau (pg/mL) | 433 (213) | 320 (133) | 533 (222) | **<.0001** | **<.0001** |
| p-Tau181 (pg/mL) | 66.4 (30.2) | 51.3 (15.1) | 79 (33.8) | **<.0001** | **<.0001** |
| Blood biomarkers | All | Aβ- | Aβ+ | P | P\$ |
| Fasting glycemia (mmol/L) | 5.4 (1.1) | 5.4 (1.1) | 5.4 (1.2) | .99 | .71 |
| Triglycerides (mmol/L) | 1.17 (0.6) | 1.18 (0.48) | 1.15 (0.69) | .70 | .62 |
| Cholesterol total (mmol/L) | 5.5 (1.2) | 5.5 (1.3) | 5.5 (1.1) | .78 | .29 |
| Cholesterol LDL (mmol/L) | 1.7 (0.5) | 1.8 (0.5) | 1.7 (0.5) | .58 | .81 |
| Cholesterol HDL (mmol/L) | 3.2 (1) | 3.3 (1.1) | 3.2 (0.9) | .92 | .21 |
| Prealbumin (mg/dL) | 28.4 (6.4) | 28.5 (7.6) | 28.3 (5.1) | .77 | .77 |
| Albumin (g/L) | 40.1 (4.3) | 39.9 (3.6) | 40.2 (4.9) | .60 | .38 |
| Creatinine (μmol/L) | 79.0 (23.0) | 80.1 (24.9) | 78.2 (21.5) | .54 | .76 |
| eGFR (mL/min/1.73m<sup>2</sup>) | 76.9 (14.7) | 76.9 (15.5) | 77.5 (15.5) | .62 | .45 |
| Plasma P-tau181(pg/mL) | 3.3 (1.5) | 2.6 (1.4) | 3.9 (1.4) | **<.0001** | **<.0001** |

Abbreviations: MCI: mild cognitive impairment; MCI, APOE: apolipoprotein E; BMI, body mass index; eGFR, estimated glomerular filtration rate; MMSE, Mini-Mental State Examination; R+L, right+left.
P: Comparison between the three groups, by ANOVA or $\chi^2$ ; P\$: comparison between the three groups by linear regression adjusted for age, sex, and the presence of the APOE ε4 allele; % (number) were used to describe categorical variables, mean ± standard deviation for continuous variables.

### Comparison between MCI converters and non-converters: P-Tau predicts conversion to AD

At baseline, the MCI converters to dementia were older, had a lower MMSE score, were more often APOE ε4 carriers and had more severe hippocampal atrophy (Table 1). As lumbar puncture was optional in our cohort, cerebrospinal fluid values were available in 214 subjects. CSF Aβ1-40, Aβ1-42, Tau and p-Tau181 values were highly differential between MCI that converted or not. We next turned our attention to the efficacy of markers in the blood, as these samples were available from all participants. We have previously described differential levels of plasma amyloid biomarkers in the BALTAZAR cohort ^6^. Here we found plasma P-tau181 increased significantly, on average by 30%, between MCI non-converters at 2.9 [SD 1.4] pg/mL and converters at 3.8 [SD 1.5] pg/mL (p<0.0001). Plasma P-Tau remained significant after adjustment for age, sex, and APOE ε4 status (Table 1). No difference was observed between MCI converters and non-converters for metabolic or renal function blood biomarkers: fasting blood glucose (glycemia), triglycerides, cholesterol (total, HDL, LDL), prealbumin, albumin and creatinine or glomerular filtration rate (eGFR).

### Comparison between Aβ+ and Aβ-patients: Tau predicts amyloid status

The amyloid status, Aβ+ corresponding to the (A+) ATN classification ^3^, was defined based on the CSF Aβ1-42/Aβ1-40 ratio ^23^. Almost half of the MCI participants had a lumbar puncture and 117 of them were Aβ+ and 97 were Aβ-. At baseline, Aβ+ patients were older, had a lower MMSE score and were more often APOE ε4 carriers. However, unlike MCI converters, Aβ+ patients did not have a lower hippocampal volume (Table *2*). CSF Aβ1-42, Tau and p-Tau181 values were also highly differential between Aβ+ and Aβ-patients. Plasma P-tau181 was significantly higher on average by 50% in Aβ+ than in Aβ-(3.9 [SD 1.4] vs. 2.6 [SD 1.4] pg/mL, p<0.0001). This difference remained significant after adjustment for age, sex, and APOE ε4 status (Table *2*).

### P-tau181 improves predictive power of age, sex, APOEε4 status and MMSE for MCI conversion and amyloid status detection

Using a logistic regression approach with conversion as a dependent variable, and age, sex, APOEε4 status and MMSE as independent variables, it was possible to predict conversion (P <.0001) with an AUC of the model fit of 0.691 [95%CI=0.655-0.741]. The addition of plasma P-tau181 resulted in a significant increase of the AUC to 0.744 [95%CI=0.702-0.784] (Suppl Table 3). The added value of plasma P-tau181 was further documented by computing the net reclassification improvement of the two models. This revealed a 12.8% improvement in patient classification between MCI converters and non-converters due to plasma P-tau181. Since blood biomarkers are intended to replace CSF biomarkers, we compared the respective values of plasma and CSF P-tau181 for amyloid status detection in the sub-cohort where patients had undergone a lumbar puncture. The addition in the model of plasma or CSF P-tau181 resulted in a significant increase of the AUC from 0.786 [95%CI=0.723-0.84] for age, sex, APOEε4 status and MMSE to 0.849 [95%CI=0.792-0.895] and 0.857 [95%CI=0.801-0.902], respectively (P[difference]=0.00075 and 0.0002). AUCs obtained by the addition of plasma or CSF P-tau181 for conversion or amyloid status detection (0.750 and 0.752, respectively) were not different (P[difference]=.81). Importantly, when creatinine or eGFR were associated with P-tau181 using logistic regression, they did not give in better performance models.

### Association of plasma P-tau181 with other biomarkers and cohort characteristics

The relationships and correlations between plasma P-tau181 concentration and the other biomarkers and cohort characteristics were analysed after splitting the population by tertile (Table 3). Age, body mass index (BMI) and APO ε4 were significantly different between tertile. Patient conversion rate was also clearly correlated with plasma P-tau181, with values of 16.4%, 26.1% and 47.8% in the 1^st^, 2^nd^ and 3^rd^ tertile, respectively. Distribution of Aβ+ patients was also greatly increased along with tertile, a relationship that was confirmed by the high correlation observed between plasma P-tau181 and CSF Aβ1-42/Aβ1-40 use to define the Aβ+ status (correlation coefficient = -0.4428, p<0.0001). eGFR decreased (p=0.017) while creatinine very significantly increased (p<.0001) in the higher plasma P-tau181 tertiles. All these results remained significant after adjustment for age, sex, and APOE ε4 status (Table 3). The relationship between plasma P-tau181 and MCI conversion was further documented by plotting the Kaplan–Meier curve of conversion to dementia according to the tertiles (Figure 1A). A very significant overall difference was observed (Log rank P<.0001) and the hazard ratio (HR) between the 1^st^ and the 3^rd^ tertile was 3.8 [95%CI=2.5-5.8].

**Table 3:**
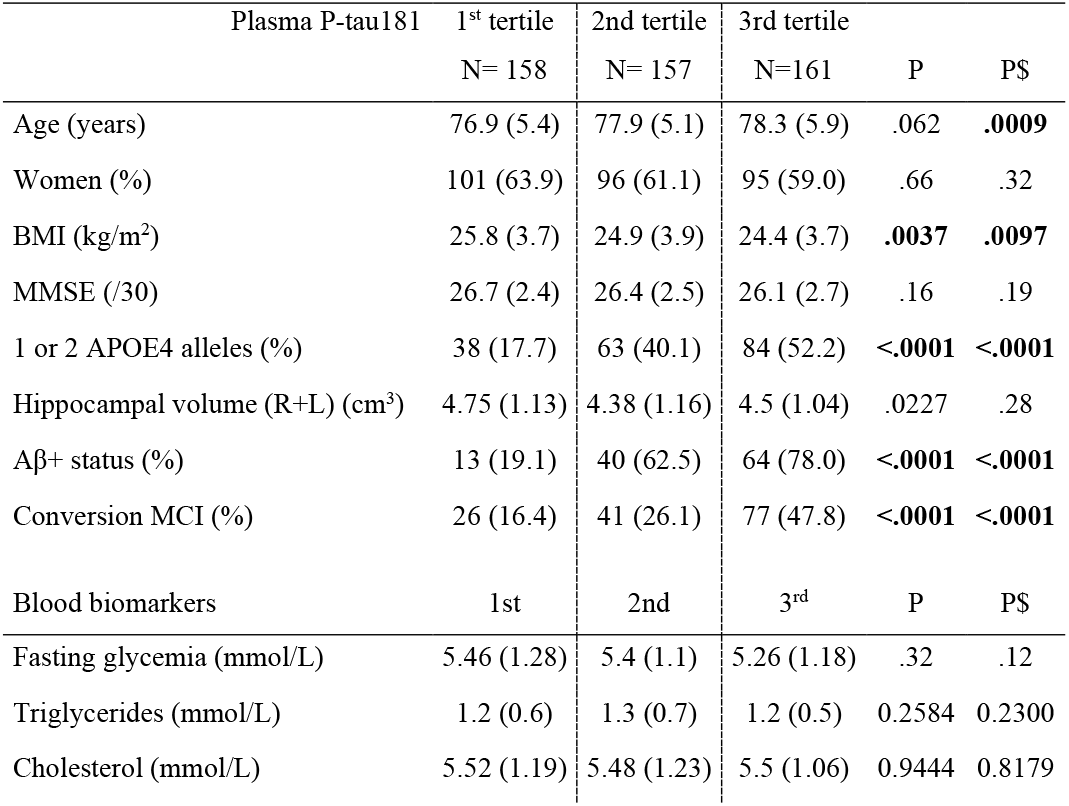
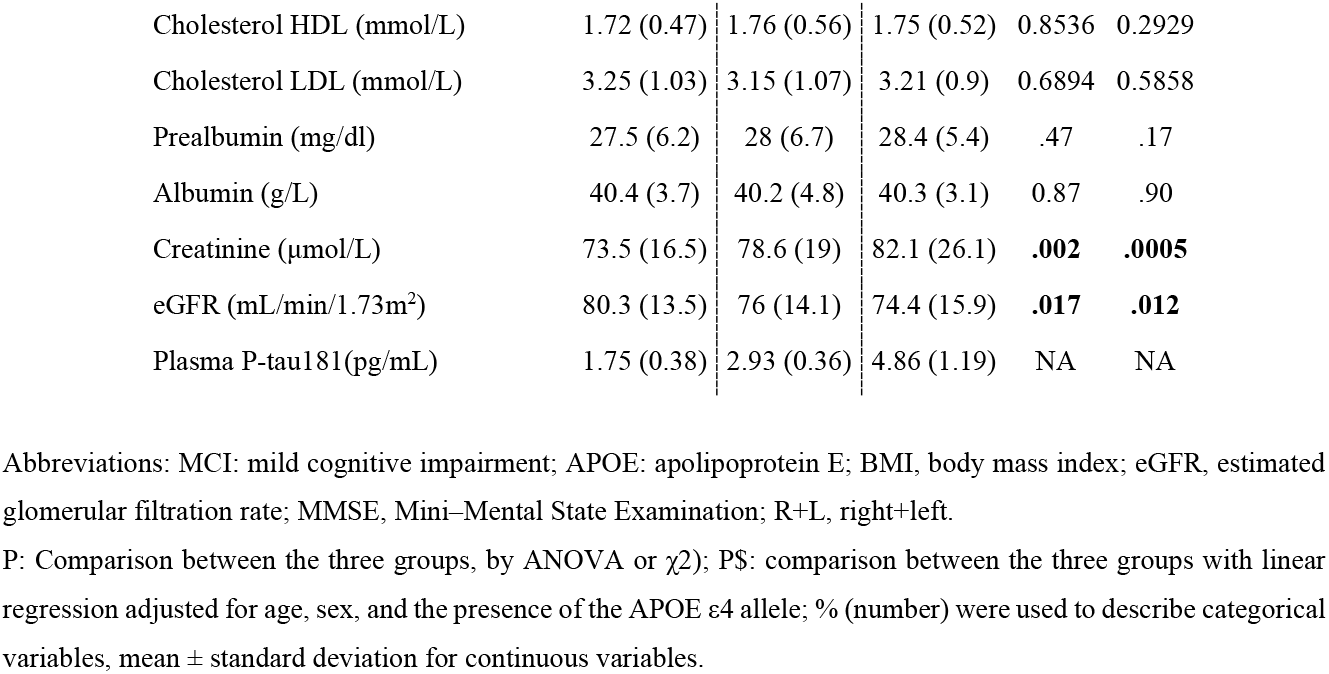
Characteristics in the different P-tau181 tertiles.

**Figure 1.**
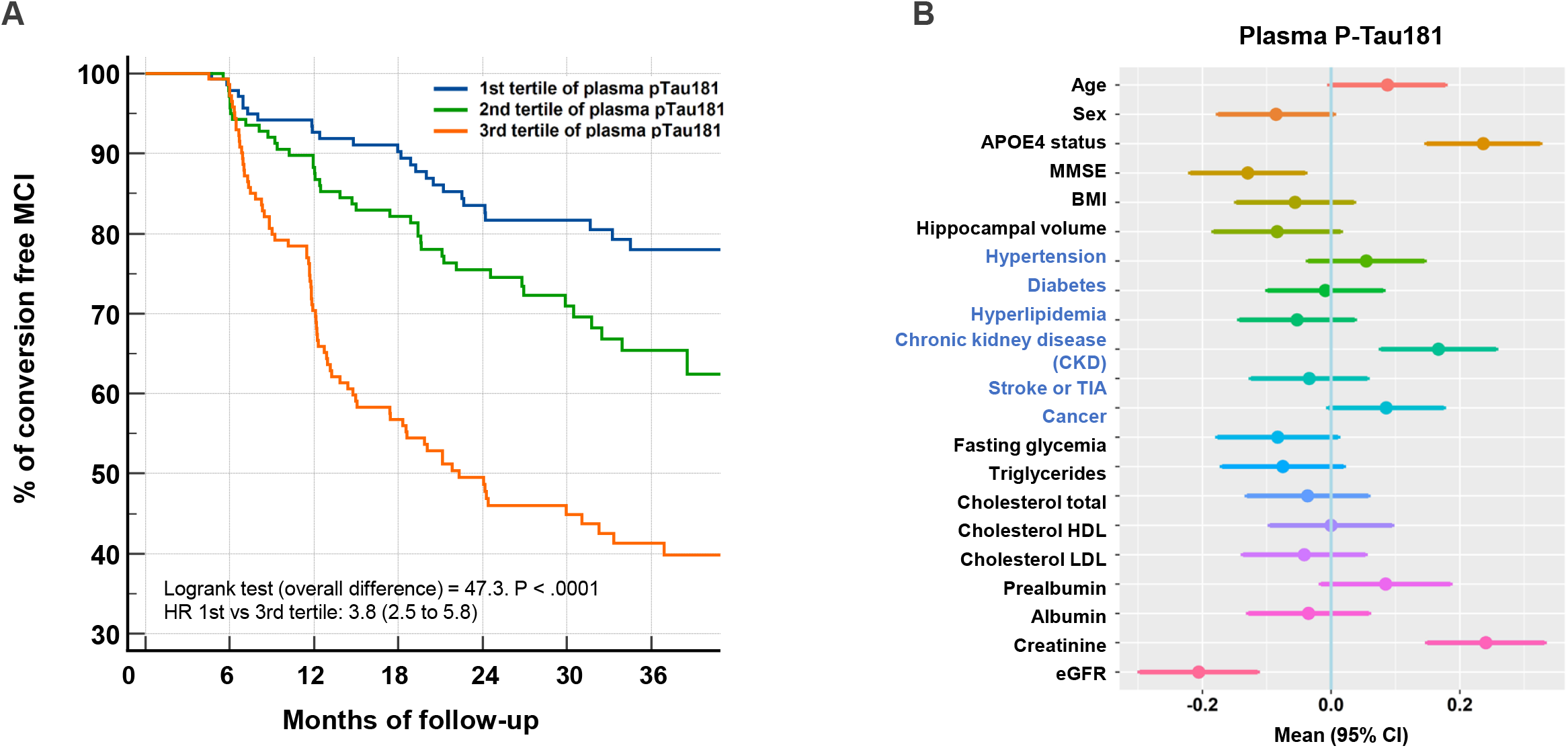
***Panel A: Kaplan–Meier curve of conversion to dementia according to the tertiles of plasma P-tau181 in MCI subjects*** Abbreviations: HR, hazard ratio; MCI, mild cognitive impairment. ***Panel B: Associations between multiple factors and plasma P-tau181 concentrations*** Forest plots of associations between demographic, comorbidities (in blue) and biological variables and plasma P-tau181, using linear regression. Means and 95% confidence intervals (CIs) are provided. Z-scores are used to compare the factors between them. Abbreviations: TIA, transient ischemic attack; APOE, apolipoprotein E; BMI, body mass index; eGFR, estimated glomerular filtration rate; MMSE, Mini–Mental State Examination.

### Impact of comorbidities and covariates on P-tau181 concentration and diagnostic performance

The relationship between plasma P-tau181 concentration and comorbidities, demographic factors and biological information collected at baseline in the BALTAZAR cohort was investigated using a linear regression approach. We identified APOE status, creatinine and eGFR as strongly connected to plasma P-tau181 (Figure 1B). To a lesser degree, age and BMI also affected plasma P-tau181levels. Among comorbidities we tested, chronic kidney disease (CKD) appeared strongly linked to P-taul181 levels. All these results remained very similar after adjustment for age, sex, and APOE ε4 status. To further evaluate the impact of these covariates, we plotted the correlation between creatinine, eGFR, BMI and age in the population stratified by amyloid status (Figure 2A-D). The correlation in the BALTAZAR subpopulation with lumbar puncture (n=214) remained significant only for eGFR and creatinine (Suppl Table 2). We observed higher values of plasma P-tau181 in the Aβ+ population. P-tau181 levels were also correlated with low and high values of eGFR and creatinine. To confirm this observation, we stratified the population based on tertile of creatinine or eGFR levels (Figure 2EF, Table 4). The analysis of variance confirmed that the mean level of plasma P-tau181 was globally differential among both creatinine or eGFR tertiles (p<0.001). Finally, to evaluate the impact on P-tau181 on the detection of amyloid status, we computed the ROC curves and determined best cutpoints and corresponding performance for Aβ+ (Table 4).

**Table 4:** Performance of plasma P-tau181 for Aβ+ detection with regard to renal function (creatinine and eGFR)

| Population with CSF biomarkers available | Total | Creatinine < 68<br>1st tertile | Creatinine 69-82<br>2nd tertile | Creatinine ≥ 82<br>3rd tertile |
| --- | --- | --- | --- | --- |
| **Creatinine** (μmol/L) | 74 [71-77] | 61 [57-64] | 73 [71-80] | 95 [84-111] |
| Plasma P-tau 181 (pg/mL) | 3.0 [2.8-3.4] | 2.3 [1.8-3.3] | 2.9 [2.5-3.9] | 3.7 [2.7-5.2] |
| Plasma P-tau 181 in Aβ- | 2.2 [1.9-2.4] | 1.9 [1.5-2.4] | 2.3 [1.6-2.7] | 2.9 [1.9-4.0] |
| Plasma P-tau 181 in Aβ+ | 3.7 [2.8-4.5] | 3.1 [2.2-4.3] | 3.6 [2.9-4.0] | 4.2 [3.6-5.4] |
| AUC Plasma P-tau 181 Aβ+ | 0.783 | 0.763 | 0.872 | 0.733 |
| Plasma P-tau 181 cutpoints (pg/mL) | 2.77 | 2.31 | 2.77 | 3.52 |
| **Sensitivity** (%) | 80.9 | 75.0 | 89.5 | 80.0 |
| **Specificity** (%) | 69.6 | 73.0 | 80.0 | 70.0 |
| Population with CSF biomarkers available | Total | eGFR < 74<br>1st tertile | eGFR 74-86<br>2nd tertile | eGFR > 86<br>3rd tertile |
| **eGFR** (mL/min/1.73m<sup>2</sup>) | 79 [68-89] | 64 [54-69] | 79 [76-84] | 91 [89-93] |
| Plasma P-tau 181 (pg/mL) | 3.0 [2.1-3.9] | 3.5 [2.4-4.2] | 3.1 [2.1-3.9] | 2.7 [1.8-3.9] |
| Plasma P-tau 181 in Aβ- | 2.2 [1.7-3.0] | 2.7 [1.7-3.4] | 2.0 [1.7-2.6] | 2.0 [1.6-2.9] |
| Plasma P-tau 181 in Aβ+ | 3.7 [2.8-4.5] | 3.8 [3.1-5.4] | 3.8 [2.8-4.5] | 3.3 [2.7-4.2] |
| AUC Plasma P-tau 181 Aβ+ | 0.783 | 0.763 | 0.872 | 0.733 |
| Plasma P-tau 181 cutpoints (pg/mL) | 2.77 | 2.81 | 2.71 | 2.44 |
| **Sensitivity** (%) | 80.9 | 86.4 | 84.2 | 78.8 |
| **Specificity** (%) | 69.6 | 60.0 | 77.8 | 68.6 |

Abbreviations: CSF: cerebrospinal fluid; AUC: area under the ROC curve; eGFR, estimated glomerular filtration rate. Values of creatinine and plasma P-tau181 are express as median [25-75 percentile]. Cutpoints correspond to the best Youden index on the ROC curves.

**Figure 2.**
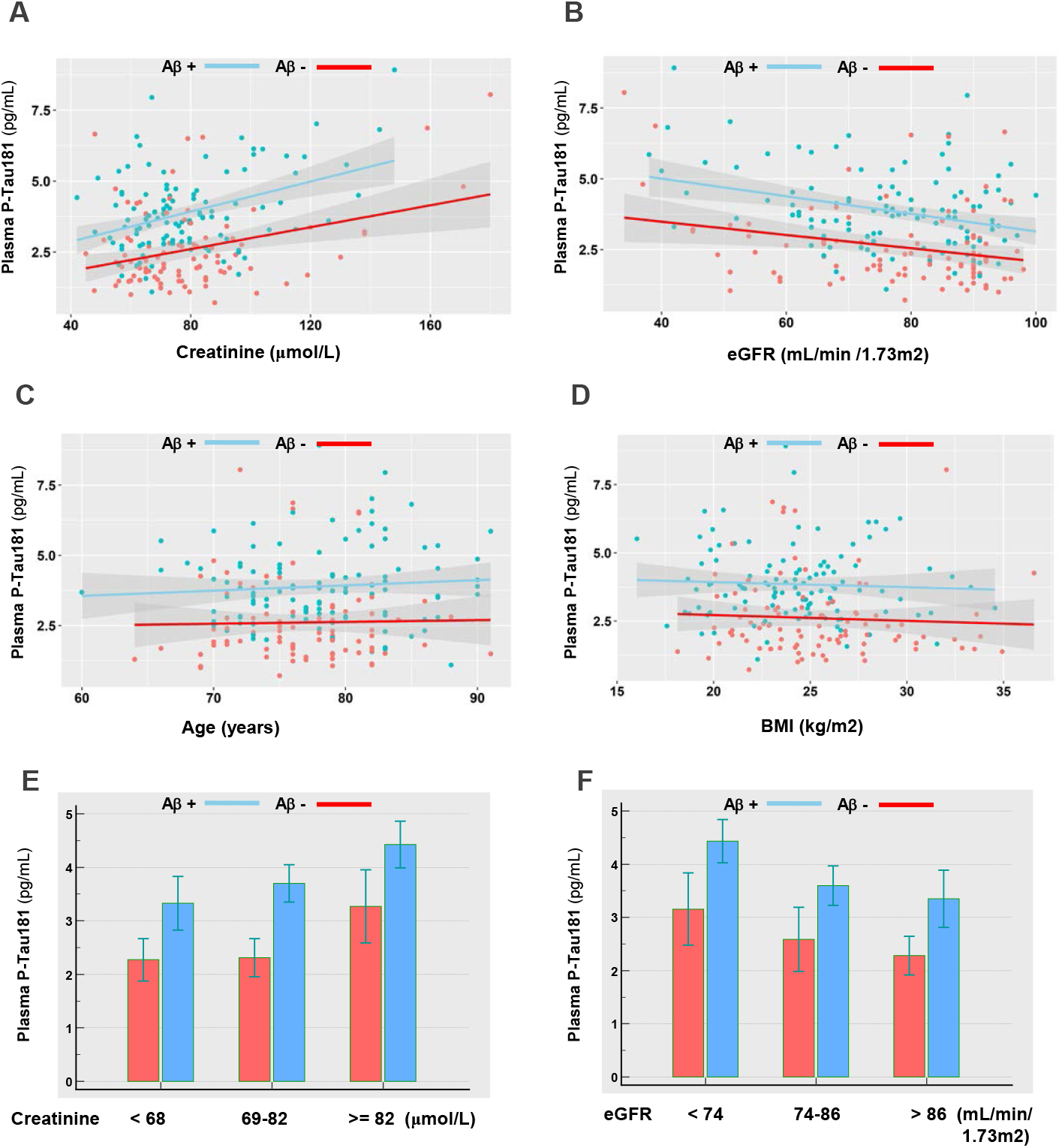
***Panels A-D: Correlation between plasma P-tau181 and creatinine, eGFR, age and BMI in the amyloid negative and positive populations.*** ***Panels E,F: Levels of plasma P-tau181 in the amyloid negative and positive populations, by tertiles of creatinine or eGFR*** Concentrations of plasma P-tau181 were significantly different between amyloid negative and positive patients in all cases, as tested using a Mann-Whitney test (P<0.05). Abbreviations: BMI, body mass index; eGFR, estimated glomerular filtration rate (unit: mL/min/1.73m^2^).

## Discussion

Here we present results from a large-scale multicenter prospective longitudinal cohort of clinically defined mild cognitive impairment (MCI) participants, referred to memory center, with a follow-up of three years. Our principal finding is that patients who convert to dementia have 30% higher levels of plasma P-tau181 independently of age, sex or APOE ε4. Importantly, 48% of MCI participants among the highest tertile of plasma P-tau181 (>3.61) converted to dementia and thus had a fourfold higher risk. In addition, patients in the first P-Tau(181) tertile (i.e., with a value ≤2.32 pg/mL) have a conversion rate of 19.8% over a 3-year period. It is likely that combining P-Tau(181) with other blood biomarkers such as plasma amyloid peptides could improve this prediction. This information is valuable for patient management and for using therapeutic strategies to prevent progression. In this MCI population, plasma P-tau181 also predicted amyloid status (based on the CSF Aβ1-42/Aβ1-40 ratio), with Aβ+ patients having 50% higher P-tau181 levels than their Aβ-counterparts.

One important element that raises the interest of using this plasma biomarker in the future is its added value above that of just using a combination of age, sex, APOEε4 status and MMSE. It is noteworthy that adding plasma P-tau181 significantly improved the detection of both Aβ+ patients and MCI converters. Even more striking is that this added value of plasma P-tau181 was equivalent to that of CSF P-tau181. This finding will impact future clinical use of the approach, as it might avoid the need for lumbar puncture. The capacity of plasma P-tau181 to detect Aβ+ patients as well as AD and MCI when compared to control and to other diseases has been described ^8 9 11-13 24-27^. However, the only previous other large study focusing on MCI conversion was that of Karikari et al. ^11^ who observed that baseline concentrations of plasma P-tau181 accurately predicted future dementia and Aβ+ status (as defined by PET). As well as validating this previous study, our study has the added value of the biological data collected in the BALTAZAR cohort. These include metabolic blood biomarkers: fasting glycemia, triglycerides, cholesterol (total, HDL, LDL), prealbumin, albumin, creatinine and eGFR, which can be used to monitor diabetes, cardiovascular risk, nutritional status or kidney function. None of these factors were differential, either in comparing MCI converters to non-converters, or when comparing Aβ+ and Aβ-patients. However, when we investigated factors influencing P-Tau181 level, by comparing tertiles or through a linear regression method, we first identified age and BMI as cofounding factors. These two factors have previously been associated with P-Tau181, as well as with other blood biomarkers like neurofilaments ^28^. Age increases both plasma and CSF values of neurodegenerative biomarkers like total tau ^29^ yet to be determined reasons. For BMI, a likely relevant factor is the dilution of neuronal biomarkers in the blood volume.

Among the comorbidities that were associated with P-tau181, CKD was the most differential. This association with CKD has already been reported in recent studies ^30 31^. However, in the BALTAZAR cohort we have access to the clinical chemistry profile realized on the same plasma sample used for P-tau181 measurement. We thus noted that P-tau181 correlates with markers of kidney function: creatinine and eGFR. Adding these parameters improved the ability of P-Tau181 to detect Aβ+ patients, whereas adding age and BMI did not. Strikingly P-Tau181 and creatinine stratify together irrespective of other variables. Namely, in situations with increased creatinine (>=82 μmol/L) or low eGFR (<74 ml/min/1.73m^2^), indicating a moderate impaired kidney function, levels of P-Tau181 were increased in both Aβ+ and Aβ-patients, as well as in MCI patients converting or not to dementia.

A major suggestion of our study is tailoring the clinical cutpoints of P-Tau181 to renal function. We advocate minimizing the false detection of a pathological situation in patients by always combining plasma P-Tau181 with an assessment of renal function, e.g., through creatinine measurement and GFR estimation. This recommendation should be confirmed for other P-Tau isoforms (P-Tau217, P-Tau231) measured by immunoassay ^25^ or mass spectrometry ^32^. We cannot exclude at this stage that altered renal function may also contribute in some way to the progression of AD^33^. Indeed, this hypothesis is supported by the difference in creatine level between naMCI and aMCI population. To understand the relationship between renal function and P-Tau levels, it’s clearance by the kidney will therefore have to be studied in more detail. Of note, only very small amounts of this biomarker were detected in the free form or associated with exosome in urine ^34 35^.

The present study has some limitations. To increase the likelihood of conversion to AD we excluded participants with Lewy Body, Parkinson, frontotemporal or vascular MCI disorders. Therefore 77% of subjects had amnestic MCI and 30% of participants developed dementia which in 95% of cases was represented by probable AD. Amyloid status was available in only a part of the population, since the BALTAZAR study focused on conversion, and it was defined using CSF biomarkers rather than with PET amyloid.

The main strengths of the study lie in the large sample size of MCI participants that are well described, the controlled pre-analytical conditions, the centralized plasma P-tau181 analyses and the availability of clinical chemistry analyte measurement realized in the same sample tube.

## Conclusion

This study of our well-characterized population confirms the clinical relevance of plasma P-tau181 for the detection of amyloid status, which is important for risk assessment, patient management and inclusion in clinical trials. We also demonstrate the strong predictive value of this blood biomarker for the prognosis of MCI patients, thus addressing an important medical need in memory centers. The question remains as to the use of blood biomarkers as a screening tool in patients without cognitive impairment who have risk factors and may benefit most from preventive strategies, and/or as triage tests in patients with early symptoms for whom future investigations, including imaging and spinal tap, are being considered. Finally, we identified and quantified the impact of renal function, assessed by creatinine levels and GFR estimation, on P-tau181 blood levels. These measures are an easy and standardizable way to provide essential information about kidney function and thus to optimize interpretation of results in routine clinical practice.

## Data Availability

Data and informed consent form are available upon request after publication (APHP, Paris). Requests will be considered by each study investigators based on the information provided by the requester regarding the study and analysis plan. If the use is appropriate, a data sharing agreement will be put in place before a fully de-identified version of the dataset including the data dictionary used for analysis with individual participant data is made available.

## Statements

### Contributors

S.L., J.S.V. and O.H. take responsibility for the integrity of the data and the accuracy of the data analysis.

Concept and design: O.H., S.B., A.G., S S-M., S.L.

Acquisition, analysis, or interpretation of data: All authors.

Drafting of the manuscript: S.L.

Critical revision of the manuscript for important intellectual content: All authors.

Statistical analysis: S.L., J.S.V.

Obtained funding: O.H., S.B., A.G., S S-M., S.L.

All authors had full access to the data and contributed to revision and editing of the manuscript.

### Data availability statement

Data and informed consent form are available upon request after publication (APHP, Paris). Requests will be considered by each study investigator based on the information provided by the requester regarding the study and analysis plan. If the use is appropriate, a data sharing agreement will be put in place before distributing a fully de-identified version of the dataset, including the data dictionary used for analysis with individual participant data.

### Ethical appoval

All participants gave written informed consent to be part of the study. The BALTAZAR study was approved by the Paris ethics committee (CPP Ile de France IV Saint-Louis Hospital).

