## Supplementary material for "Plasma phosphorylated tau 181 predicts amyloid status and conversion to dementia stage dependent on renal function" (from another version of this preprint: version 2, 2022-10-11)

***Suppl Table 1: Characteristics of participants and plasma biomarkers at baseline according to the cognitive status***

| Patient characteristics | All MCI N=476 | aMCI N=365 | naMCI N=111 | P |
|---|---|---|---|---|
| Age (years) | 77·7 (5·5) | 77·6 (5·5) | 78·1 (5·4) | ·40 |
| Women (%) | 292 (61·4) | 210 (57·4) | 82 (73·9) | **·0019** |
| BMI (kg/m2) | 25 (3·8) | 25·1 (3·8) | 24·6 (3·7) | ·21 |
| 1 or 2 APOE4 alleles (%) | 185 (39·8) | 158 (43·3) | 27 (24·3) | **·0003** |
| MMSE (/30) | 26·4 (2·5) | 26·1 (2·6) | 27·4 (2·1) | **<·0001** |
| Hippocampal volume (R+L) (cm3) | 4·55 (1·12) | 4·43 (1·16) | 4·96 (0·86) | **<·0001** |
| CSF biomarkers | All MCI | aMCI | naMCI | P |
| Aβ1-40 (pg/mL) | 7434 (2241) | 7525 (2258) | 7103 (2167) | ·25 |
| Aβ1-42 (pg/mL) | 766 (384) | 744 (394) | 848 (335) | ·07 |
| Aβ1-42/Aβ1-40 | 0·104 (0·045) | 0·1 (0·046) | 0·12 (0·038) | **·0026** |
| Tau (pg/mL) | 578 (254) | 486 (247) | 322 (152) | **<·0001** |
| p-Tau181 (pg/mL) | 76·7 (30·2) | 74·3 (33·9) | 56·1 (19·0) | **<·0001** |
| Blood biomarkers | All MCI | aMCI | naMCI | P |
| Fasting glycemia (mmol/L) | 5·37 (1·19) | 5·43 (1·3) | 5·19 (0·7) | **·018** |
| Triglycerides (mmol/L) | 1,21 (0,59) | 1,24 (0,6) | 1,14 (0,53) | 0,1409 |
| Cholesterol (mmol/L) | 5,5 (1,16) | 5,45 (1,15) | 5,65 (1,18) | 0,1434 |
| Cholesterol HDL (mmol/L) | 1,74 (0,52) | 1,7 (0,52) | 1,87 (0,49) | 0,0030 |
| Cholesterol LDL (mmol/L) | 3,2 (1) | 3,18 (1) | 3,27 (1,01) | 0,4754 |
| Creatinine (μmol/L) | 78·2 (21·3) | 79·3 (22·3) | 74·5 (17·2) | **·022** |
| Prealbumin (mg/dL) | 27·6 (5·4) | 28·2 (6·4) | 27·2 (5·9) | ·14 |
| Albumin (g/L) | 40·3 (3·9) | 40·5 (4·1) | 39·8 (3·1) | ·10 |
| eGFR (mL/min/1·73m2) | 76·9 (14·7) | 76·7 (15·2) | 77·4 (13·1) | ·68 |
| Plasma P-tau181(pg/mL) | 3·19 (1·49) | 3·29 (1·5) | 2·87 (1·42) | **·0083** |

Abbreviations: MCI: mild cognitive impairment; aMCI, amnestic MCI; naMCI, nonamnestic MCI; APOE: apolipoprotein E; BMI, body mass index; eGFR, estimated glomerular filtration rate; MMSE, Mini–Mental State Examination; R+L, right+left;

P: Comparison between the aMCI and naMCI, with Student t test or $\chi 2$;
% (number) were used to describe categorical variables. mean ± standard deviation for continuous variables.
CSF biomarkers were available only in 179 and 49 patients with aMCI and naMCI, respectively.

***Suppl Table 2: Correlation between plasma P-tau181 and covariates in the Aβ- and Aβ+ populations***

| Amyloid status | Plasma P-tau181 vs. | Age | BMI | eGFR | Creatinine |
|---|---|---|---|---|---|
| Aβ- | Correlation coefficient | 0·065 | -0·079 | -0·256 | 0·337 |
| | Significance Level P | 0·5301 | 0·4429 | **0·0139** | **0·001** |
| Aβ+ | Correlation coefficient | 0·075 | -0·009 | -0·341 | 0·41 |
| | Significance Level P | 0·4203 | 0·9254 | **0·0002** | **<0·0001** |

***Suppl Table 3: Added value of plasma or CSF P-tau181 in combination with age, sex, APOE4 and MMSE for MCI conversion and Aβ+ detection***

| Population / comparison | Logistic regression with | AUC | 95% CI | AUCs comparison | NRI |
|---|---|---|---|---|---|
| All study population MCI non-converters vs MCI converters | Age, sex, APOE4 and MMSE | 0·691 | 0·655 to 0·741 | Delong test **< ·0001** | 12·8% |
| | + Plasma P-tau181 | 0·744 | 0·702 to 0·784 | | |
| Patients with CSF biomarkers MCI non-converters vs MCI converters | Age, sex, APOE4 and MMSE | 0·694 | 0·629 to 0·753 | Delong test | |
| | + Plasma P-tau181 | 0·750 | 0·687 to 0·805 | **< ·0001** | 17·5% |
| | + CSF P-tau181 | 0·752 | 0·690 to 0·807 | **< ·0001** | 17·5% |
| Patients with CSF biomarkers Aβ- vs Aβ+ | Age. sex. APOE4 and MMSE | 0·786 | 0·723 to 0·840 | Delong test | |
| | + Plasma P-tau181 | 0·849 | 0·792 to 0·895 | ·0075 | 13·7% |
| | + CSF P-tau181 | 0·857 | 0·801 to 0·902 | ·0002 | 15·9% |

Abbreviations: CSF: cerebrospinal fluid; MCI: mild cognitive impairment; APOE: apolipoprotein E; MMSE, Mini–Mental State Examination; AUC: area under the ROC curve; CI: confidence interval; NRI: net reclassification improvement. The AUCs of the combination Age+Sex+APOE4+MMSE+Plasma P-tau181 and Age+Sex+APOE4+MMSE+CSF P-tau181 were not significantly different for amyloid or conversion detection, as tested using Delong test (P=·81).

BALTAZAR study group

Olivier Hanon [1], Frédéric Blanc [2], Yasmina Boudali [1], Audrey Gabelle [3], Jacques Touchon [3], Marie-Laure Seux [1], Hermine Lenoir [1], Catherine Bayle [1], Stéphanie Bombois [4], Christine Delmaire [4], Xavier Delbeuck [5], Florence Moulin [1], Emmanuelle Duron [6], Florence Latour [7], Matthieu Plichart [1], Sophie Pichierri [8], Galdric Orvoën [1], Evelyne Galbrun [9], Giovanni Castelnovo [10], Lisette Volpe-Gillot [11], Florien Labourée [1], Pascaline Cassagnaud [12], Claire Paquet [13], Françoise Lala [14], Bruno Vellas [14], Julien Dumurgier [13], Anne-Sophie Rigaud [1], Christine Perret-Guillaume [15], Eliana Alonso [16], Foucaud du Boisgueheneuc [17], Laurence Hugonot-Diener [1], Adeline Rollin-Sillaire [12], Olivier Martinaud [18], Clémence Boully [1], Yann Spivac [19], Agnès Devendeville [20], Joël Belmin [21], Philippe-Henri Robert [22], Thierry Dantoine [23], Laure Caillard [1], David Wallon [24], Didier Hannequin [18], Nathalie Sastre [14], Sophie Haffen [25], Anna Kearney-Schwartz [15], Jean-Luc Novella [26], Vincent Deramecourt [12], Valérie Chauvire [27], Gabiel Abitbol [1], Nathalie Schwald [19], Caroline Hommet [28], François Sellal [29], Marie-Ange Cariot [16], Mohamed Abdellaoui [30], Sarah Benisty [31], Salim Gherabli [1], Pierre Anthony [29], Frédéric Bloch [32], Nathalie Charasz [1], Sophie Chauvelier [1], Jean-Yves Gaubert [1], Guillaume Sacco [22], Olivier Guerin [22], Jacques Boddaert [33], Marc Paccalin [17], Marie-Anne Mackowiak [12], Marie-Thérèse Rabus [9], Valérie Gissot [34], Athanase Benetos [15], Candice Picard [20], Céline Guillemaud [35], Gilles Berrut [8], Claire Gervais [22], Jaques Hugon [13], Jean-Marc Michel [29], JeanPhilippe David [19], Marion Paulin [12], Pierre-Jean Ousset [14], Pierre Vandel [36], Sylvie Pariel [21], Vincent Camus [37], Anne Chawakilian [1], Léna Kermanac'h [1], Anne-Cécile Troussiere [12], Cécile Adam [23], Diane Dupuy [20], Elena Paillaud [16], Hélène Briault [9], Isabelle Saulnier [38], Karl Mondon [37], Marie-Agnès Picat [23], Marie Laurent [16], Olivier Godefroy [20], Rezki Daheb [16], Stéphanie Libercier [29], Djamila Krabchi [1], Marie Chupin [39], Luc Buée [42], JeanSébastien Vidal [1], Edouard Chaussade [1], Sylvain Lehmann [40], Bernadette Allinquant [41], Susanna Schraen-Maschke [42].

Affiliations of the BALATAZAR group

[1] Université de Paris, EA 4468, APHP, Hopital Broca, Memory Resource and Research Centre of de Paris-Broca-Ile de France, F-75013 Paris, France.
[2] Université de Strasbourg, CHRU de Strasbourg, Memory Resource and Research Centre of Strasbourg/Colmar, French National Centre for Scientific Research (CNRS), ICube Laboratory and Fédération de Médecine Translationnelle de Strasbourg (FMTS), Team Imagerie Multimodale Intégrative en Santé (IMIS)/Neurocrypto, F-67000 Strasbourg, France.
[3] Université de Montpellier, CHRU Montpellier, Memory Research and Resources center, department of Neurology, Inserm INM NeuroPEPs team, excellence center of neurodegenerative disorders, F-34000 Montpellier, France.
[4] Univ. Lille, CHU Lille, Memory Resource and Research Centre of Lille, Department of Neurology, Inserm U1171, Degenerative and Vascular Cognitive Disorders, F-59000 Lille, France.
[5] Univ. Lille, Inserm U1171 Degenerative and Vascular Cognitive Disorders, F-59000 Lille, France.
[6] Université Paris-Saclay, APHP, Hôpital Paul Brousse, département de gériatrie, UVSQ, Inserm, CESP, Team MOODS, F-94800 Villejuif, France.

[7] Centre Hospitalier de la Côte Basque, Department of Gerontology, F-64100 Bayonne, France.
[8] Université de Nantes, EA 4334 Movement-Interactions-Performance, CHU Nantes, Memory Research Resource Center of Nantes, F-44000 Nantes, France.
[9] Sorbonne Universités, AP-HP, Centre Hospitalier Émile-Roux, Department of Gérontology 2, F-94450 Limeil-Brévannes, France.
[10] CHU de Nimes, Neurology Department, F-30000 Nimes, France.
[11] Hôpital Léopold Bellan, Memory Clinic, F-75014 Paris, France.
[12] Univ. Lille, CHU de Lille, Memory Resource and Research Centre of Lille, Department of Neurology, F-59000 Lille, France.
[13] Université de Paris, APHP, Groupe Hospitalier Saint Louis-LariboisièreFernand Widal, Memory Resource and Research Centre of Paris Nord-Ile de France, F-75010 Paris, France.
[14] Université de Toulouse III, CHU La Grave-Casselardit, Memory Resource and Research Centre of Midi-Pyrénées, F-31300 Toulouse, France.
[15] Université de Lorraine, CHRU de Nancy, Memory Resource and Research Centre of Lorraine, F-54000 Nancy, France.
[16] AP-HP, HEGP, Service de Gériatrie, F-75015, Paris, France.
[17] CHU de Poitiers, Memory Resource and Research Centre of Poitiers, F-86000 Poitiers, France.
[18] CHU Charles Nicolle, Memory Resource and Research Centre of HauteNormandie, F-76000 Rouen, France.
[19] APHP, Centre Hospitalier Émile-Roux, Department of Gérontology 1, F-94450 Limeil-Brévannes, France.
[20] CHU d'Amiens-Picardie, Memory Resource and Research Centre of Amiens Picardie, F-80000 Amiens, France.
[21] APHP, Hôpitaux Universitaires Pitie-Salpêtrière-Charles Foix, Service de Gériatrie Ambulatoire, F-75013 Paris, France.
[22] CHU de Nice, Memory Research Resource Center of PACA Est, F-06100 Nice, France.
[23] CHU de Limoges, Memory Research Resource Center of Limoges, F-87000 Limoges, France.
[24] Université de Rouen Normandie, CHU Charles Nicolle, Memory Resource and Research Centre of Haute-Normandie, Inserm U1079, IRIB, F-76000 Rouen, France.
[25] CHU de Besançon, Memory Resource and Research Centre of BesançonFranche-Comté, F-25000 Besançon, France.
[26] Université de Reims Champagne Ardenne, EA 3797, CHU de Reims, Memory Resource and Research Centre of Champagne Ardenne, F-51100 Reims, France.
[27] CHU d'Angers, Memory Resource and Research Centre of Angers, F-49000 Angers, France.
[28] CHRU de Tours, Memory Resource and Research Centre of Tours, F-37000 Tours, France.
[29] Université de Strasbourg, CHRU de Strasbourg, Memory Resource and Research Centre of Strasbourg/Colmar, Inserm U-118, F-67000 Strasbourg, France.
[30] Univ Paris Est Creteil, EA 4391 Excitabilité Nerveuse et Thérapeutique, CHU Henri Mondor, Department of Neurology, F-94000 Créteil, France.
[31] Fondation ophtalmologique Adolphe de Rothschild, Department of Neurology, F-75019 Paris, France.
[32] CHU d'Amiens-Picardie, Department of Gerontology, F-80000 Amiens, France.

[33] Sorbonne Universités, APHP, Hôpitaux Universitaires Pitie-Salpêtrière-Charles Foix, Memory Resource and Research Centre, Centre des Maladies Cognitives et Comportementales IM2A, Inserm UMR 8256, F-75013 Paris, France.
[34] Université François-Rabelais de Tours, CHRU de Tours, Memory Resource and Research Centre of Tours, Inserm CIC 1415, F-37000 Tours, France.
[35] Sorbonne Universités, APHP, Hôpitaux Universitaires Pitie-Salpêtrière-Charles Foix, Memory Resource and Research Centre, Centre des Maladies Cognitives et Comportementales IM2A, F-75013 Paris, France.
[36] Université Bourgogne Franche-Comté, EA 481 Neuroscience, IFR 133, CHU de Besançon, Memory Resource and Research Centre of BesançonFranche-Comté, F-25000 Besançon, France.
[37] Université François-Rabelais de Tours, CHRU de Tours, Memory Resource and Research Centre of Tours, Inserm U930 Imagerie et Cerveau, F-37000 Tours, France.
[38] Université de Limoges, EA 6310 HAVAE, CHU de Limoges, Memory Research Resource Center of Limoges, F-87000 Limoges, France.
[39] Université Paris-Saclay, Neurospin, CEA, cati-neuroimaging.com, CATI Multicenter Neuroimaging Platform, F-91190 Gif-sur-Yvette, France.
[40] LBPC-PPC, Université de Montpellier, INM INSERM, IRMB CHU de Montpellier, Montpellier, France.
[41] Université de Paris, Inserm UMR-S 894, F-75014 Paris, France.
[42] Univ. Lille, Inserm, CHU Lille, U1172-LilNCog, LiCEND, LabEx DISTALZ, F-59000 Lille, France.
